# Antimicrobial stewardship programs and antibiotic use in Africa: a Systemic review and meta-analysis protocol

**DOI:** 10.1101/2025.03.24.25324562

**Authors:** Ammas Siraj, Dawit Abrham

## Abstract

**Background:** Antimicrobial resistance (AMR) is a global health crisis, particularly acute in Africa where infectious disease burdens are high and resources limited. Inappropriate antibiotic use, driven by factors like easy access and inadequate diagnostics, fuels AMR. Antimicrobial stewardship programs (ASPs) offer a key intervention, but their implementation and impact in Africa remain poorly understood. This systematic review and meta-analysis evaluates the association between ASPs and antibiotic use in Africa, aiming to inform policy and practice by synthesizing existing evidence and identifying research gaps.

**Methods:** This systematic review and meta-analyses will include all original articles that will examine antimicrobial stewardship programs (ASPs) in Africa. Non-English articles, qualitative studies, editorials, comment pieces and expert opinion and review articles will be excluded. Initially limited search of MEDLINE (PubMed) and CINAHL (EBSCO) will be conducted to identify articles on the topic. The text words contained in the titles and abstracts of relevant articles, and the index terms used to describe the articles will be used to develop a full search strategy for reporting the name of the relevant databases/information sources. Data extraction will be performed by 2 independent reviewers on the articles selected for inclusion. The different characteristics of studies will be extracted. The quality of included studies will be reported using Effective Public Health Practice Project (EPHPP) quality assessment tool. Summary of the extracted data will be presented in tabular format and graphs.

**Discussion:** This protocol is expected to convey pooled evidence on association of targeted ASPs with antimicrobial consumption. Evidence from this review will be used to tackle current global problem related with anti-microbial resistance. Therefore, our review will call for government and non-government interventions in reducing current challenge of global issue related with anti-microbial resistance in resource limited settings like Africa.

**Systematic review registration number:** PROSPERO CRD420251003018.

## Introduction

Antimicrobial resistance (AMR) is a grave threat global public health, posing a major risk to the effective treatment of infectious diseases [1]. Globally, 4.95 million deaths were associated with bacterial AMR in 2019, with the greatest number of deaths per head of population in Africa [2]. The African continent faces a particularly acute challenge due to a convergence of factors, including high rates of infectious diseases, limited access to diagnostic facilities, and widespread inappropriate antibiotic use [3, 4, 5]. This situation leads to the rapid emergence and dissemination of AMR, threatening public health outcomes [3, 6].

The inappropriate use of antibiotics, driven by factors such as over-prescription, self-medication, and inadequate infection control practices, is a major driver of AMR [7, 8, 9]. Uncertainly, AMR is a global challenge that affects every continent and country regardless of their development profile. However, in Africa, these issues are exacerbated by fragile healthcare systems, inadequate resources, and a dearth of strong surveillance mechanisms [5, 10]. Consequently, infections that were once easily treatable are becoming increasingly difficult and costly to manage, leading to increased morbidity and mortality [11].

Antimicrobial stewardship programs (ASPs) are recognized as a vital strategy to optimize antibiotic use and mitigate the spread of AMR [12]. These programs aim to promote the rational use of antibiotics through various interventions, including education, guidelines, and monitoring [13]. While the importance of ASPs is well-established, their implementation and effectiveness in the African context remain variable [14].

Existing research suggests that implementing ASPs in resource-limited settings can be challenging due to factors such as limited infrastructure, inadequate staffing, and cultural beliefs [15, 16]. However, there is also evidence that tailored ASPs can lead to significant improvements in antibiotic prescribing practices and reductions in AMR [16, 17].

Therefore, a comprehensive synthesis of the available evidence is needed to assess the impact of ASPs on antibiotic use in Africa. This systematic review and meta-analysis aimed at quantifying the effect of ASPs on antibiotic consumption and prescribing patterns in African healthcare settings, and it helps to identify factors that influence the effectiveness of ASPs in this region. Moreover, it provides evidence-based recommendations for the implementation of context-effective ASPs in Africa.

Per our preliminary screening of the literature in PubMed and Google Scholar, and a search of registered systematic review databases such as PROSPERO and Open Science Frameworks (OSF, currently there are no systematic review and meta-analysis or published protocols investigating the impact of ASPs on antibiotic use in Africa.

## Study objectives

### General objective

The objective of this study is to identify the association between antimicrobial stewardship programs (ASPs) and antibiotic use in Africa

### Specific objectives

To examine the effect of ASPs on antibiotic consumption and prescribing patterns in African healthcare settings.

To determine effectiveness of different ASP interventions on antibiotic use in Africa

To identify factors that influence the association between ASPs and antibiotic use in Africa.

### Review questions

The question of this review include:

What is the impact of ASPs on antibiotic and prescribing patterns in African healthcare settings?

What is the effectiveness of different ASP interventions on antibiotic use in Africa?

What are the factors that influence the effectiveness of ASPs in this African region?

### Inclusion criteria

#### Participants

Studies conducted in any African country assess the impact of antimicrobial stewardship programs on antibiotic use across healthcare settings. Patients receiving antibiotic treatment in African healthcare facilities and healthcare professionals (doctors, nurses, pharmacists) involved in antibiotic prescribing and administration in African healthcare facilities and healthcare facilities themselves, as the units where ASPs are implemented would participants of this systematic review and meta-analysis.

#### Intervention(s)

This systematic review and meta-analyses will include all original articles that will examine antimicrobial stewardship programs (ASPs) in Africa. ASPs may include various interventions; educational programs for healthcare professionals, implementation of antibiotic usage guidelines, audit and feedback mechanisms and Restriction of certain antibiotics.

#### Outcomes

This review will consider studies that include the following outcomes: The main outcome is pooled magnitude of antibiotics consumption, which is calculated as proportion of patients receiving an antibiotic prescription and defined daily doses per 100 patient-days. The secondary outcome will be the pooled association of targeted ASPs with antimicrobial consumption, which is calculated in the form of two-by-two tables and reported using Odds ratio.

#### Phenomena of interest

This review will include original studies conducted to assess the impact of antimicrobial stewardship programs on antibiotic use across Healthcare settings in Africa.

#### Context

All original articles conducted in Healthcare settings in Africa will be included in the systematic review and meta-analyses.

#### Types of research syntheses

This systematic review and meta-analyses will consider original studies of all different designs. The goal of EPHPP systematic review and meta-analyses is to reveal pooled magnitude of effectiveness [18, 19]. The review will also include qualitative syntheses of research articles.

## Methods

The planned review of systematic review and meta-analyses will be carried out using the EPHPP systematic review and meta-analyses methodology [18]. The title for this review was registered on PROSPERO and obtained registration ID CRD420251003018.

### Search strategy

Comprehensive search including both published and unpublished research syntheses as per JBI Methodological recommendation [20, 21] will be sought after using the search strategies. In this review, a three-step search methodology will be used. First, an initial limited search of MEDLINE (PubMed) and CINAHL (EBSCO) will be undertaken to identify articles on the topic. The text words contained in the titles and abstracts of relevant articles, and the index terms used to describe the articles will be used to develop a full search strategy for reporting the name of the relevant databases/information sources. The search strategy, including all identified keywords and index terms, will be adapted for each included database and/or information source. The reference list of all included sources of evidence will be screened for additional studies.

Studies published in English language will be considered. All original articles conducted in Africa on the impact of antimicrobial stewardship programs on antibiotic use across healthcare settings irrespective of year of publication fulfilling inclusion criteria will be included.

### Information sources

The databases will be searched for this review include MEDLINE (PubMed), Embase, CINAHL and Web of Science. To capture unpublished reviews, additional searches will be conducted using Google Scholar and institutional websites such as WHO website, CDC website, National Ministries of Health websites of African countries, African universities repositories.

### Study selection

The studies retrieved through data base searching and other sources will be screened for relevance, and those identified as being potentially eligible will be fully assessed against the inclusion/exclusion criteria, and selected or rejected, as appropriate. The duplicate will be removed by using the Endnote software. Critical appraisal will be conducted to assess the internal (systematic error level) and external validity (generalizability) of the studies, and to reduce the risks of biases. The mean scores of two authors will be taken into account in coming to a final decision regarding study quality, and studies with scores greater than or equal to half the required points will be included in the data analysis. A third reviewer will resolve any discrepancies arising between the two authors during the assessment process. The quality of the included studies will be evaluated by two reviewers independently using the Joanna Briggs Institute Critical Appraisal Tools (for observational studies).

### Assessment of methodological quality

Selected syntheses will be critically appraised by two independent reviewers (AS and DA) for methodological quality in the review using Effective Public Health Practice Project (EPHPP) quality assessment tool to assess 6 domains of quality: (1) selection bias, (2) design, (3) confounders, (4) blinding, (5) data collection methods, and (6) withdrawal and dropouts. EPHPP is a widely used assessment tool for quantitative studies designed for systematic literature reviews of effectiveness studies. [18]. Authors of papers will be contacted to request missing or additional data for clarification, where required. Any disagreements that arise between the reviewers will be resolved through discussion, or with a third reviewer. The results of the critical appraisal will be reported in narrative form and in a table. All syntheses, regardless of the results of their methodological quality, will undergo data extraction and synthesis.

### Data collection

The review began in April 2025, and the official literature search will be finished by the end of June 2025. Quantitative data will then be extracted from the papers selected for inclusion using the standardized JBI data extraction tool. The authors will extract important data related to the study characteristics (the region and the study area, the first author, the year of publication, the study design, the population characteristics, and the sample size) and the outcomes of interest (the effect size data, including the antimicrobial stewardship program, and the antibiotic use). Any disagreements that arose between the reviewers were resolved through discussion or with a third reviewer. Authors of papers were contacted to request missing or additional data, where required. Any papers with missed information after two contacts of authors of the included studies will be excluded if no response obtained at after the second contact.

### Data summary

Data from included studies will be extracted using tool prepared in Microsoft Excel and exported to Stata version 18.0 for analyses of the pooled estimates of primary and secondary outcome measures, as well as for subgroup analysis. Considering variation in true effect sizes across the population (clinical heterogeneity), DerSimonian and Laird’s random effects model will be applied for the analyses, with a 95% confidence level. Accordingly, the event size (proportion) will be calculated, and standard error of Logit event rate will be determined using Stata software. A quantitative synthesis will be carried out using random effects model. We will also provide a narrative synthesis of the findings, structured around the predictive factors investigated, the target population characteristics, and the type of outcome content. Summaries of the strength of the association between the prognostic risk factors and the outcomes for each study will also be provided by calculating risk ratios (for dichotomous outcomes) or standardised mean differences (for continuous outcomes).

### Assessing certainty in the outcome

The Grading of Recommendations, Assessment, Development and Evaluation (GRADE) approach for grading the certainty of evidence will be followed and a Summary of Findings (SoF) will be created using GRADEpro GDT [22] (McMaster University, ON, Canada). We will use the GRADE approach to determine how confident we are in the evidence from the reviews we include. The GRADE approach evaluates four aspects: the quality and design of the original studies, how consistent their results are, and how relevant they are to our question.

We will rate the evidence as high, moderate, low, or very low quality. We will also summarize the findings for the four outcomes that our review focuses on: Antibiotics consumption, prescribing practice, ASPs effectiveness, broad spectrum antibiotics prescribing [21]. This will be undertaken by two independent reviewers (AS and DA) at the outcome level. Any disagreements that arise between the reviewers will be resolved through discussion or with a third reviewer.

Authors of papers were contacted to request missing or additional data for clarification, where required through two emails request. The SoF will present the following information where appropriate: absolute risks for the treatment and control, estimates of relative risk, and a ranking of the quality of the evidence based on the risk of bias, directness, heterogeneity, precision and risk of publication bias of the review results. The outcomes reported in the SoF will be antibiotics use in Africa and ASPs as primary and secondary outcomes respectively.

### Assessing confidence in the synthesized findings

The final synthesized findings will be graded according to the ConQual approach for establishing confidence in the output of qualitative research synthesis and presented in a Summary of Findings [23]. The Summary of Findings includes the major elements of the review and details how the ConQual score is developed. Included in the Summary of Findings will be the title, population, phenomena of interest, and context for the specific review. Each synthesized finding from the review will then be presented, along with the type of research informing it, the score for dependability and credibility, and the overall ConQual score.

## Discussion

Antimicrobial resistance (AMR) is a global health threat closely linked to the inappropriate use of antibiotics, which can diminish the effectiveness of antibiotics. It is a significant and growing problem in Africa, with high rates of antibiotic resistance reported. Inappropriate antibiotic use is a major driver of AMR, making ASPs crucial. Many African countries face resource limitations, which can hinder the implementation and sustainability of ASPs. Lack of trained personnel, limited laboratory capacity, and inadequate funding are common challenges. The prevalence and effectiveness of ASPs vary significantly across African countries due to differences in healthcare systems, resources, and policies. ASPs have the potential to significantly reduce inappropriate antibiotic use in Africa. Strategies such as antibiotic formulary restrictions, pre-authorization requirements, and prospective audit and feedback can promote rational antibiotic prescribing.

Despite the potential benefits, ASP implementation faces challenges, including: Lack of awareness and understanding of ASP principles, Resistance from healthcare providers, Weak healthcare infrastructure. Effective ASPs in Africa require context-specific approaches that consider local epidemiology, resource availability, and cultural factors. The WHO, and various other organizations are pushing for the implementation of ASP’s across the African continent. There is a need for more research to evaluate the effectiveness of ASPs in different African settings. Improved surveillance of antibiotic use and resistance is essential to monitor the impact of ASPs. While ASPs are recognized as a vital tool in combating AMR in Africa, their successful implementation requires addressing significant challenges. Efforts are being made to promote ASPs across the continent, but sustained commitment and investment are needed.

As far as researchers’ knowledge is concerned this systematic review and meta-analyses is of its first kind in Africa on ASPs and antibiotics use. The review will focus to identify the overall pooled magnitude of antibiotics consumption, and pooled association of targeted ASPs with antimicrobial consumption in Africa. The result will helps to provide evidence-based recommendations for the implementation of effective ASPs in Africa, which might be base to design policies and strategies to fight against anti-microbial and irrational use of antibiotics across different healthcare settings in Africa.

All published and unpublished original article in Africa setting will be considered better estimating the overall pooled magnitude of antibiotics consumption, and pooled association of targeted ASPs with antimicrobial consumption in Africa. Hence, this systematic review and meta-analyses is required to reveal the overall pooled magnitude of antibiotics consumption, and pooled association of targeted ASPs with antimicrobial consumption in Africa. It is also essential opportunity with study to state the status of ASPs and its impacts on antibiotics use in Africa. The study will also significantly important to embrace the factors that influence the association between ASPs and antibiotic use in Africa, to enable tailored intervention will be designed for each factor.

The result from this systematic review and meta-analysis will attract the attentions of government and partners for ASPs which is serving as driving engine to fight against anti-microbial resistance. Furthermore, it serves for policy makers by providing evidence on the impacts of ASPs on antimicrobial use to design context specific intervention to optimize antimicrobial use and halt misuse of antimicrobial. Parallel to this, it will supplement evidence for government to develop guidelines, policies and proclamations for use of antimicrobial use which will play pivotal role in preventing anti-microbial resistance. The result form this review will essential for appropriate implementation of across all healthcare settings in Africa. It can be taken as a benchmark for future researchers undertaking their interventional and other high standard studies to optimize ASPs and reduce anti-microbial resistances.

All efforts will be made to disseminate the evidence to scientific community through different means like present the work on different workshops, conference and publication on internationally reputable scientific journal. However, the two might be limitation of the review like related with language, and limitation of study area. This can cause bias on the outcome. Therefore, the finding of this review should use with caution to the other areas than within Africa.

## Conclusion

The finding will used as a basic information for understanding the impact of ASP on antibiotics use in Africa. Furthermore, this review will aid for design of context specific tool to implement ASP in Africa as antimicrobial resistance is one of the global concerns currently. In addition, the result will explore importance of tailored ASP implementation in prevention of anti-microbial resistance-Global concerns of our time. Provide evidence-based recommendations for the implementation of effective ASPs in Africa.

## Data Availability

No datasets were generated or analysed during the current study. All relevant data from this study will be made available upon study completion.

## Acknowledgments

The authors acknowledge all involved in this study.

## Author contributions

**Conceptualization:** Ammas Siraj

**Data curation:** Dawit Abrham

**Formal analysis:** Ammas Siraj and Dawit Abrham

## Funding acquisition

**Investigation:** Dawit Abrham

**Methodology:** Ammas Siraj and Dawit Abrham

**Project administration:** Ammas Siraj and Dawit Abrham

**Resources:** Ammas Siraj and Dawit Abrham

**Software:** Ammas Siraj

**Supervision**: Ammas Siraj

**Validation:** Ammas Siraj and Dawit Abrham

**Visualization:** Dawit Abrham

**Writing** – original draft: Ammas Siraj and Dawit Abrham

**Writing** – review & editing: Ammas Siraj and Dawit Abrham

## Notes

### Competing Interest Statement

The authors have declared no competing interest.

### Funding Statement

The author(s) received no specific funding for this work.

### Author Declarations

This is a systematic review and meta-analyses protocol which doesn't need ethical approval.

## References

1. O’neill J. Antimicrobial resistance: tackling a crisis for the health and wealth of nations; 2014:4–5. Available from: https://amr-review.org/sites/default/files/AMR%20Review%20Paper%20-%20Tackling%20a%20crisis%20for%20the%20health%20and%20wealth%20of%20nations_1.pdf. Accessed January 15, 2025.

2. Murray C, Ikuta K, Sharara F, et al. Global burden of bacterial antimicrobial resistance in 2019: a systematic analysis. Lancet. 2022;399(10325):629–655. doi: 10.1016/S0140-6736(21)02724-0

3. Ayukekbong JA, Ntemgwa M, Atabe AN. The threat of antimicrobial resistance in developing countries: causes and control strategies. Antimicrob Resist Infect Control. 2017;6(1):47. doi: 10.1186/s13756-017-0208.

4. Asha Ripanda, Mwemezi J. Rwiza, Elias Charles Nyanza et al Ecological consequences of antibiotics pollution in sub-Saharan Africa: Understanding sources, pathways, and potential implications, Emerging Contaminants, Volume 11, Issue 2, 2025, 100475, ISSN 2405-6650, 10.1016/j

5. Gulumbe BH, Haruna UA, Almazan J, Ibrahim IH, Faggo AA, Bazata AY. Combating the menace of antimicrobial resistance in Africa: a review on stewardship, surveillance and diagnostic strategies. Biol Proced Online. 2022 Nov 23;24(1):19. doi: 10.1186/s12575-022-00182

6. Godman B, Egwuenu A, Wesangula E, et al. Tackling antimicrobial resistance across sub-Saharan Africa: current challenges and implications for the future. Expert Opin Drug Saf. 2022;21(8):1089–1111. doi: 10.1080/14740338.2022.2106368.

7. Ndaki, P.M., Mwanga, J.R., Mushi, M.F. et al. Drivers of inappropriate use of antibiotics among community members in low- and middle-income countries: a systematic review of qualitative studies. BMC Public Health 25, 705 (2025). 10.1186/s12889-025-21553-6

8. Gashaw, T., Yadeta, T.A., Weldegebreal, F. et al. The global prevalence of antibiotic self-medication among the adult population: systematic review and meta-analysis. Syst Rev 14, 49 (2025). 10.1186/s13643-025-02783-6

9. Anant Nepal, Delia Hendrie, Linda A. Selvey, Suzanne Robinson, Factors influencing the inappropriate use of antibiotics in the Rupandehi district of Nepal, 25 August 2020. 10.1002/hpm.3061

10. Otaigbe II, Elikwu CJ. Drivers of inappropriate antibiotic use in low- and middle-income countries. JAC Antimicrob Resist. 2023 May 31;5(3):dlad062. doi: 10.1093/jacamr/dlad062. PMID: 37265987; PMCID: PMC10230568.

11. Sirwan Khalid Ahmed, Safin Hussein, Karzan Qurbani, Radhwan Hussein Ibrahim, Abdulmalik Fareeq, Kochr Ali Mahmood, Mona Gamal Mohamed, Antimicrobial resistance: Impacts, challenges, and future prospects, Journal of Medicine, Surgery, and Public Health, Volume 2, 2024, 100081, ISSN 2949-916X,10.1016/j.glmedi.2024.100081.

12. Jishna Shrestha; Farah Zahra; Preston Cannady, Jr. Antimicrobial Stewardship, June 20, 2023. https://www.ncbi.nlm.nih.gov/books/NBK572068/

13. Nathwani, D., Varghese, D., Stephens, J. et al. Value of hospital antimicrobial stewardship programs [ASPs]: a systematic review. Antimicrob Resist Infect Control 8, 35 (2019). 10.1186/s13756-019-0471-0

14. Chukwu EE, Abuh D, Idigbe IE, Osuolale KA, Chuka-Ebene V, Awoderu O, Audu RA, Ogunsola FT. Implementation of antimicrobial stewardship programs: A study of prescribers’ perspective of facilitators and barriers. PLoS One. 2024 Jan 19;19(1):e0297472. doi: 10.1371/journal.pone.0297472.

15. Rolfe R Jr, Kwobah C, Muro F, Ruwanpathirana A, Lyamuya F, Bodinayake C, Nagahawatte A, Piyasiri B, Sheng T, Bollinger J, Zhang C, Ostbye T, Ali S, Drew R, Kussin P, Anderson DJ, Woods CW, Watt MH, Mmbaga BT, Tillekeratne LG. Barriers to implementing antimicrobial stewardship programs in three low- and middle-income country tertiary care settings: findings from a multi-site qualitative study. Antimicrob Resist Infect Control. 2021 Mar 25;10(1):60. doi: 10.1186/s13756-021-00929-4.

16. Asrat Agalu Abejew, Gizachew Yismaw Wubetu, Teferi Gedif Fenta, Assessment of challenges and opportunities in antibiotic stewardship program implementation in Northwest Ethiopia, Heliyon, Volume 10, Issue 11, 2024, e32663, ISSN 2405-8440, 10.1016/j.heliyon.2024.e32663.

17. Harun, M.G.D., Sumon, S.A., Hasan, I. et al. Barriers, facilitators, perceptions and impact of interventions in implementing antimicrobial stewardship programs in hospitals of low-middle and middle countries: a scoping review. Antimicrob Resist Infect Control 13, 8 (2024). 10.1186/s13756-024-01369-6

18. Effective Public Health Practice Project. Quality assessment tool for quantitative studies. Accessed December 21, 2022. https://www.ephpp.ca/PDF/Quality%20Assessment%20Tool_2010_2.pdf

19. Page, M.J., McKenzie, J.E., Bossuyt, P.M. et al. The PRISMA 2020 statement: an updated guideline for reporting systematic reviews. Syst Rev 10, 89 (2021). 10.1186/s13643-021-01626-4

20. Cooper, C., Booth, A., Varley-Campbell, J. et al. Defining the process to literature searching in systematic reviews: a literature review of guidance and supporting studies. BMC Med Res Methodol 18, 85 (2018). 10.1186/s12874-018-0545-3

21. homas BH, Ciliska D, Dobbins M, Micucci S. A process for systematically reviewing the literature: providing the research evidence for public health nursing interventions. Worldviews Evid Based Nurs. 2004;1(3):176–184.

22. Guyatt G, Oxman AD, Akl EA, Kunz R, Vist G, Brozek J, et al. GRADE guidelines: 1. Introduction-GRADE evidence profiles and summary of findings tables. J Clin Epidemiol. 2011;64(4):383–94.10.1016/j.jclinepi.2010.04.026

23. Munn Z, Porritt K, Lockwood C, Aromataris E, Pearson A. Establishing confidence in the output of qualitative research synthesis: the ConQual approach. BMC Med Res Methodol. 2014;14:108. 10.1186/1471-2288-14-108

